# High-flow nasal cannula vs non-invasive ventilation in acute hypoxia: Propensity score matched study

**DOI:** 10.1101/2023.09.26.23296167

**Authors:** Elizabeth S Munroe, Ina Prevalska, Madison Hyer, William J Meurer, Jarrod M Mosier, Mark A. Tidswell, Hallie C Prescott, Lai Wei, Henry Wang, Christopher M Fung

## Abstract

**RATIONALE:** The optimal treatment for early hypoxemic respiratory failure is unclear, and both high-flow nasal cannula and non-invasive ventilation are used. Determining clinically relevant outcomes for evaluating non-invasive respiratory support modalities remains a challenge.

**OBJECTIVES:** To compare the effectiveness of initial treatment with high-flow nasal cannula versus non-invasive ventilation for acute hypoxemic respiratory failure.

**METHODS:** We conducted a retrospective cohort study of patients with acute hypoxemic respiratory failure treated with high-flow nasal cannula or non-invasive ventilation within 24 hours of Emergency Department arrival (1/2018-12/2022). We matched patients 1:1 using a propensity score for odds of receiving non-invasive ventilation. The primary outcome was major adverse pulmonary events (28-day mortality, ventilator-free days, non-invasive respiratory support hours) calculated using a Win Ratio.

**MEASUREMENTS AND MAIN RESULTS:** 1,265 patients met inclusion criteria. 795 (62.8%) received high-flow oxygen and 470 (37.2%) received non-invasive ventilation. We propensity score matched 736/1,265 (58.2%) patients. There was no difference between non-invasive ventilation vs high-flow nasal cannula in 28-day mortality (17.7% vs 23.1%, p=0.08) or ventilator-free days (median [Interquartile Range]: 28 [25, 28] vs 28 [13, 28], p=0.50), but patients on non-invasive ventilation required treatment for fewer hours (median 7 vs 13, p< 0.001). Win Ratio for composite major adverse pulmonary events favored non-invasive ventilation (1.26, 95%CI 1.06-1.49, p< 0.001).

**CONCLUSIONS:** In this observational study of patients with acute hypoxemic respiratory failure, initial treatment with non-invasive ventilation was superior to high-flow nasal cannula for major pulmonary adverse events. Evaluation of composite outcomes is important in the assessment of respiratory support modalities.

## Introduction

Acute hypoxemic respiratory failure is a major cause of hospitalizations in the US.^1,2^ There is growing evidence that non-invasive respiratory support may help prevent invasive mechanical ventilation in patients with respiratory failure.^3^ Traditionally, the primary mode of non-invasive respiratory support was non-invasive positive pressure ventilation (NIV)—continuous positive airway pressure and bilevel positive airway pressure. Over the past decade, accelerated by the COVID-19 pandemic, high-flow nasal canula (HFNC) has emerged as an alternative.^4,5^ NIV and HFNC work through different mechanisms and thus have different benefits and harms. NIV improves oxygenation by increasing mean airway pressure but has the potential to deliver injurious lung volumes which may put patients at risk for patient self-induced lung injury.^3,6,7^ In contrast, HFNC provides less positive pressure ventilatory support than NIV, which may decrease the risk for self-induced lung injury and can help improve patient tolerance.^7–10^

The optimal mode of non-invasive respiratory support in acute hypoxemic respiratory failure remains unclear.^3,11^ While guidelines recommend NIV for patients with acute decompensated heart failure and chronic obstructive pulmonary disease exacerbations, these recommendations are based on comparisons of NIV to low-flow oxygen not HFNC.^3,11^ Guidelines further conclude that there is not enough evidence to make a recommendation for HFNC vs NIV in other etiologies of hypoxemic respiratory failure.^3^ The few trials that have directly compared HFNC to NIV have been limited by their focus on narrow populations.^4,12–15^ The largest trial found that for intensive care unit (ICU) patients with pure hypoxemic respiratory failure, HFNC decreased mortality compared to NIV.^13^ However, trials in different populations, e.g., COVID-19 or immunocompromised patients, have yielded conflicting results.^4,14,15^ Furthermore, the generalizability of these trials, particularly to undifferentiated hypoxemic respiratory failure in the emergency department, is limited given patients often require initiation of respiratory support before a clinical diagnosis can be made.

Additionally, many of the primary outcomes used in trials of non-invasive respiratory support, such as intubation and ventilator-free days, hinge on subjective practice decisions that limit their interpretation.^16^ Composite outcomes can help overcome the limitations of potentially subjective individual endpoints. However, traditional approaches to generating composite outcomes lead to loss of the relative importance of key variables (e.g., mortality and need for intubation are treated similarly) and have statistical limitations that prevent the combination of different types of variables (e.g., binary mortality and continuous ventilator-free days). The Win Ratio, which has been broadly applied in cardiology trials, offers a novel approach to composite outcomes, allowing the combination of different types of variables ranked by clinical relevance.^17^ The goal of this study was to compare the effectiveness of initial treatment of acute hypoxemic respiratory failure with HFNC versus NIV on a composite outcome of major adverse pulmonary events, calculated using a Win Ratio.

## Materials and Methods

### Study design and setting

This is a single-center, propensity score matched, retrospective cohort study comparing initial treatment with HFNC vs NIV in patients with acute hypoxemic respiratory failure. This study was approved by the University of Michigan (UM) Institutional Review Board on 3/14/2023 (HUM00232776) as a secondary data use, exempt study.

### Data Source

We extracted data from the electronic health record (EPIC, Epic systems, Verona, Wisconsin) via queries to our health system’s data warehouses (Clarity and Caboodle) and, for respiratory support mode data, directly from clinical flowsheets. Each variable used in our analysis, its definition and source are listed in **eTable 1**. There were no missing data for the exposure or outcome variables. Encounters with missing data on covariates in the propensity score model were excluded.

### Inclusion and Exclusion Criteria

We included patients ≥18 years-old presenting to the University of Michigan adult emergency department (ED) between 1/2018-12/2022 who received HFNC or NIV within 24 hours of ED arrival. To capture acute respiratory failure, we excluded patients who were on chronic home ventilator support, had a tracheostomy, or received non-invasive respiratory support only after extubation. We also excluded patients who had positive SARS-CoV-2 antigen test and clinically suspected COVID-19 infection. At our institution, HFNC was the predominant respiratory support mode used for patients with COVID-19 based on concerns about aerosolization and virus transmission with NIV. A single patient may have had multiple encounters during the study period; each encounter was considered an individual observation.

### Data collection and definition of respiratory support mode

We identified patients who received HFNC or NIV within 24 hours of presentation to the ED based on clinical flowsheets. At our institution, respiratory therapists and nurses record respiratory support mode and settings in a flowsheet at least once per hour and whenever mode or setting changes are made. We defined HFNC as any delivery of greater than 20 liters/minute oxygen from a heated, humidified oxygen system. We defined NIV as any non-invasive ventilation mode (continuous positive airway pressure or bilevel positive airway pressure). We classified patients who received more than one non-invasive respiratory support mode during a given hour according to the dominant mode (mode used most frequently) for the hour. For the primary analysis, we classified patients as receiving initial HFNC or NIV if they received those modes exclusively for the first two hours after initiation, to ensure stable initial group assignment.

### Study Outcomes

The primary outcome was a hierarchical composite outcome of major adverse pulmonary events: 28-day mortality, 28-day ventilator-free days, and hours spent on non-invasive respiratory support from initiation through hour 72. The primary outcome was calculated using a Win Ratio (see *Data Analysis* below for details). We also evaluated the components of the composite major adverse pulmonary events outcome individually: mortality and time to death (28-day and in-hospital), intubation rate, ventilator-free days, and non-invasive respiratory support hours.

### Data Analysis

We used a logistic regression model to calculate a propensity score for the odds of receiving NIV using pre-specified patient characteristics typically available at the time of initiation of HFNC or NIV: age, sex, body mass index, Charlson comorbidity index, history of congestive heart failure, chronic obstructive pulmonary disease (COPD), or obstructive sleep apnea, comparison of measured pCO_2_ by blood gas (venous or arterial) to expected pCO_2_ by Winter’s formula, time from ED arrival to HFNC or NIV initiation, initial lactate, initial Glasgow Coma Score, and highest day 1 sequential organ failure assessment (SOFA) score. While SOFA score is not available on ED arrival, it was included in the propensity score model because it provides a surrogate measure for severity of illness. ICD-10 diagnosis codes were recorded to understand etiologies of respiratory failure but were not included in the propensity score model, as discharge-level diagnoses are not available at the time of HFNC or NIV initiation.

Using the propensity score, we matched patients 1:1 utilizing greedy nearest neighbor matching with calipers set at 0.2 standard deviations.^18^ We used standard descriptive statistics, including standard mean differences (SMD) and Kolmogorov-Smirnov statistcs, to assess for covariate balance and distribution before and after propensity-score matching. SMD < 0.1 after matching was considered acceptable balance.

To calculate the primary outcome of major pulmonary adverse events, we used a Win Ratio, a statistical method for combining variables into a hierarichal composite outcome. Unlike typical composite outcomes that assign similar weight to outcomes regardless of their severity (i.e., including both all-cause mortality and need for repeat intervention) or take an “all-or-nothing” approach to evaluating the presence vs absence of an event, the Win Ratio accommodates mixed variable types (e.g., binary, ordinal, continuous, time-to-event) and ranks variables by clinical relevance and patient priorities. Prior literature has defined the calculation of the Win Ratio in detail.^17^ In brief, we generated all possible pairs of patients on NIV and HFNC. Pairs were sequentially compared on outcomes based on pre-determined importance: 1) 28-day mortality as a survival event, 2) 28-day ventilator-free days, and 3) non-invasive respiratory support hours from initiation to hour 72. We incorporated selective tie-breaking to best parallel clinical reasoning. For example, for two individuals who tied on mortality, we did not compare ventilator-free days or non-invasive respiratory support hours. Total wins and losses were added up across all three outcome tiers and compared to generate the Win Ratio, which was calculated using the WinRatio package (v1.0).^19^

Individual secondary outcomes were compared using Mann-Whitney U tests for continuous variables and Chi-squared tests for categorical variables. Time to death and time to intubation were analyzed using a Kaplan-Meier survival analysis. Hourly respiratory support mode and daily patient status were visualized using stacked histogram plots.

We conducted several sensitivity analyses: 1) intention-to-treat framework: inclusion of patients who crossed-over between respiratory support modes or had discontinuation of non-invasive respiratory support in the first 2 hours, 2) alternative approaches to calculating the Win Ratio.

Initial data cleaning, data exploration, and visualization was performed using Tableau (Tableau Software). Final data cleaning and all statistical analyses were performed using R (v4.2.1) in R Studio (v2022.2.3).

## Results

Between January 2018 and December 2022, 2,208/361,459 (0.6%) adult ED encounters required non-invasive respiratory support within 24 hours of ED arrival. Of these, 1,265/2,208 (57.3%) met study inclusion criteria (**Figure 1**). HFNC was the predominant mode, used in 795/1,265 (62.8%) encounters. Patients treated with NIV and HFNC had large differences in baseline congestive heart failure (78.3% vs 54.5%, SMD 0.52), pCO_2_ above expected based on Winter’s Formula (67.7% vs 33.7%, SMD 0.74), shock index (heart rate/systolic blood pressure: 0.7 vs 0.8, SMD 0.36) and lactate >4 mmol/L (9.8% vs 15.0%, SMD 0.22) (**Table 1**). The most common encounter diagnosis codes were respiratory failure not otherwise specified (89.7% vs 85.5%), followed by volume overload (54.1%) and pneumonia (48.9%) in the HFNC group and volume overload (78.1%) and COPD/asthma exacerbation (52.3%) in NIV group (**eTable 2**).

**Figure 1:**
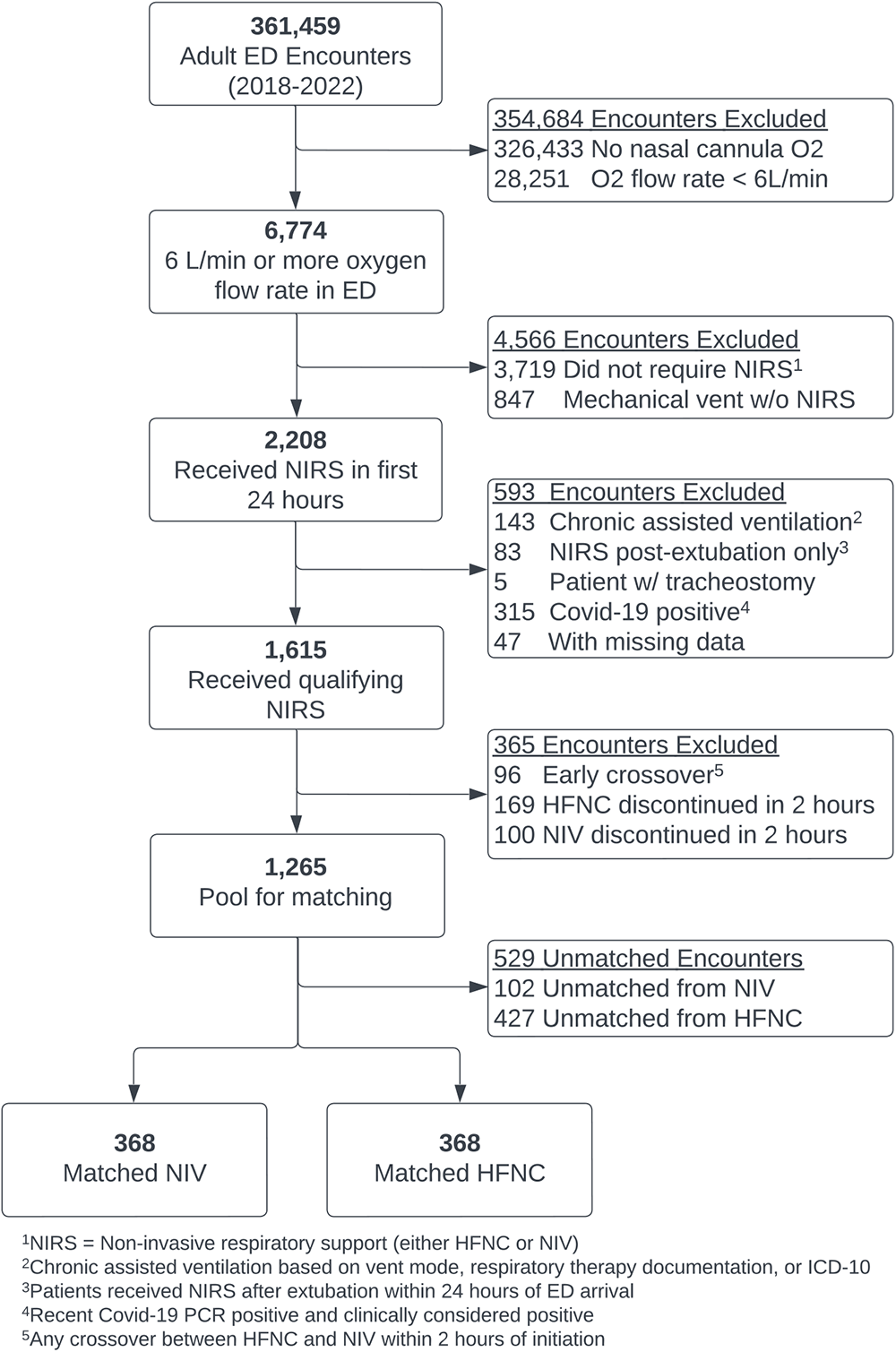
Study flow diagram. The primary analysis included N=1,265 patients in the pool for matching. A sensitivity analysis was performed for all patients receiving qualifying non-invasive respiratory support (NIRS), including early cross-over and discontinuation (N=1,615, see eTable 1). <u>Definitions</u>: ED= Emergency Department, O2= oxygen, L= liter, NIRS= non-invasive respiratory support, which includes both high-flow nasal cannula and non-invasive ventilation, vent= ventilation, HFNC= high-flow nasal canula, NIV= non-invasive ventilation, which includes continuous positive airway pressure and bilevel positive airway pressure.

**Table 1.** Baseline characteristics in the overall cohort and matched study cohort. Data are presented as median [IQR] and N(%). *Expected pCO_2_ was calculated using the first available bicarbonate from a basic metabolic panel and pCO_2_ from the first arterial or venous blood gas, comparing actual pCO_2_ to pCO_2_ that would be expected based on Winter’s formula. pCO_2_ above expected suggests superimposed respiratory alkalosis, while pCO_2_ below expected suggests superimposed respiratory acidosis. Patients were matched on calculated expected pCO_2_, not individual pH, pCO_2_, or bicarbonate values. <u>Definitions:</u> HFNC= high flow nasal cannula, NIV= non-invasive positive pressure ventilation, SMD= standard mean difference, systolic shock index= ratio of heart rate / blood pressure, SOFA= sequential organ failure assessment. Non-invasive respiratory support refers to both HFNC and NIV.

|  | Overall Cohort |  |  | Matched Cohort |  |  |
| --- | --- | --- | --- | --- | --- | --- |
|  | HFNC<br>(n=795) | NIV<br>(n=470) | SMD | HFNC<br>(n = 368) | NIV<br>(n = 368) | SMD |
| Age, year | 66.7 [55.4, 75.7] | 68.9 [59.6, 77.6] | 0.218 | 69.4 [57.1, 78.2] | 68.3 [59.0, 77.6] | 0.031 |
| Male sex | 466 (58.6) | 245 (52.1) | 0.131 | 194 (52.7) | 199 (54.1) | 0.027 |
| Body Mass Index, kg/m2 | 26.3 [22.1, 31.2] | 29.0 [23.6, 35.5] | 0.395 | 27.4 [23.1, 32.9] | 27.5 [23.0, 34.3] | 0.109 |
| Charlson Comorbidity Index | 8.0 [4.0, 14.0] | 9.0 [5.0, 14.0] | 0.154 | 8.0 [4.0, 14.0] | 8.5 [5.0, 14.0] | 0.047 |
| Individual comorbidities at baseline |  |  |  |  |  |  |
| Congestive heart failure | 433 (54.5) | 368 (78.3) | 0.521 | 264 (71.7) | 266 (72.3) | 0.012 |
| Chronic obstructive pulmonary disease | 482 (60.6) | 326 (69.4) | 0.184 | 246 (66.8) | 246 (66.8) | <0.001 |
| Obstructive sleep apnea | 160 (20.1) | 132 (28.1) | 0.187 | 86 (23.4) | 96 (26.1) | 0.063 |
| Expected pCO<sub>2</sub>* |  |  |  |  |  |  |
| pCO<sub>2</sub> at expected | 184 (23.1) | 70 (14.9) | 0.74 | 70 (19.0) | 68 (18.5) | 0.065 |
| pCO<sub>2</sub> above expected | 268 (33.7) | 318 (67.7) |  | 207 (56.2) | 218 (59.2) |  |
| pCO<sub>2</sub> below expected | 343 (43.1) | 82 (17.4) |  | 91 (24.7) | 82 (22.3) |  |
| First hours of non-invasive respiratory support | 2.0 [1.0, 7.0] | 2.0 [1.0, 4.8] | 0.272 | 2.0 [1.0, 5.0] | 2.0 [1.0, 5.0] | 0.026 |
| Systolic shock index | 0.8 [0.7, 1.0] | 0.7 [0.5, 0.9] | 0.362 | 0.7 [0.6, 0.9] | 0.7 [0.6, 0.9] | 0.020 |
| Lactate group, n(%) |  |  |  |  |  |  |
| < 2 mmol/L | 415 (52.2) | 293 (62.3) | 0.22 | 210 (57.1) | 220 (59.8) | 0.058 |
| 2-4 mmol/L | 261 (32.8) | 131 (27.9) |  | 102 (29.6) | 104 (28.3) |  |
| >4 mmol/L | 119 (15.0) | 46 (9.8) |  | 49 (13.3) | 44 (12.0) |  |
| Glasgow Coma Score | 15 [15, 15] | 15 [15, 15] | 0.106 | 15 [15, 15] | 15 [15, 15] | 0.095 |
| Highest Day 1 SOFA | 5.0 [4.0, 7.0] | 5.0 [3.0, 6.0] | 0.275 | 5.0 [4.0, 6.0] | 5.0 [4.0, 7.0] | 0.016 |

In our propensity model, (**eFigure 1**) body mass index (Odds Ratio (OR) 1.03 [per kg/m^2^ increase], 95%CI: 1.02-1.05), history of congestive heart failure (OR 2.29, 95%CI: 1.70-3.11), and pCO2 above expected by Winter’s formula (OR 3.49, 95%CI: 2.48-4.96) were associated with increased odds of treatment with NIV, while hemodynamic instability (shock index: OR 0.46 [per unit increase], 95%CI: 0.26-0.82) and higher SOFA (OR 0.91 [per point increase], 95%CI: 0.86-0.96) were associated with lower odds of receiving NIV.

We matched 736/1,265 (58.1%) eligible patients at a ratio of 1:1, including 368/795 (45.9%) on HFNC and 368/470 (78.3%) on NIV. Patient characteristics were well-balanced between groups after matching, with all SMDs <0.1 except body mass index (SMD 0.109) (**Table 1, eFigure 2**). Matched NIV patients were similar to overall NIV patients, while matched HFNC patients had higher rates of congestive heart failure, higher body mass index, and more pCO_2_ above expected by Winter’s formula than overall HFNC patients (**Table 1**).

The primary outcome was major adverse pulmonary events. We first evaluated individual components of the major adverse pulmonary events outcome (**Table 2**). There was no difference between patients treated with NIV vs HFNC in 28-day mortality (17.7% vs 23.1%, p=0.08) or ventilator-free days (median [IQR]: 28 [25,28] vs 28 [13, 28], p=0.50). However, patients treated with NIV spent significantly fewer hours on non-invasive respiratory support within the first 72 hours of initiation (median 7 vs 13 hours, p<0.001). Hourly respiratory support modes up to 72 hours post-HFNC or NIV initiation and daily patient status out to 28 days by treatment group are presented in the supplement (**eFigure 3 and 4**). Time to death by 28 days was also similar between patients treated with NIV vs HFNC (**eFigure 5**).

**Table 2.**
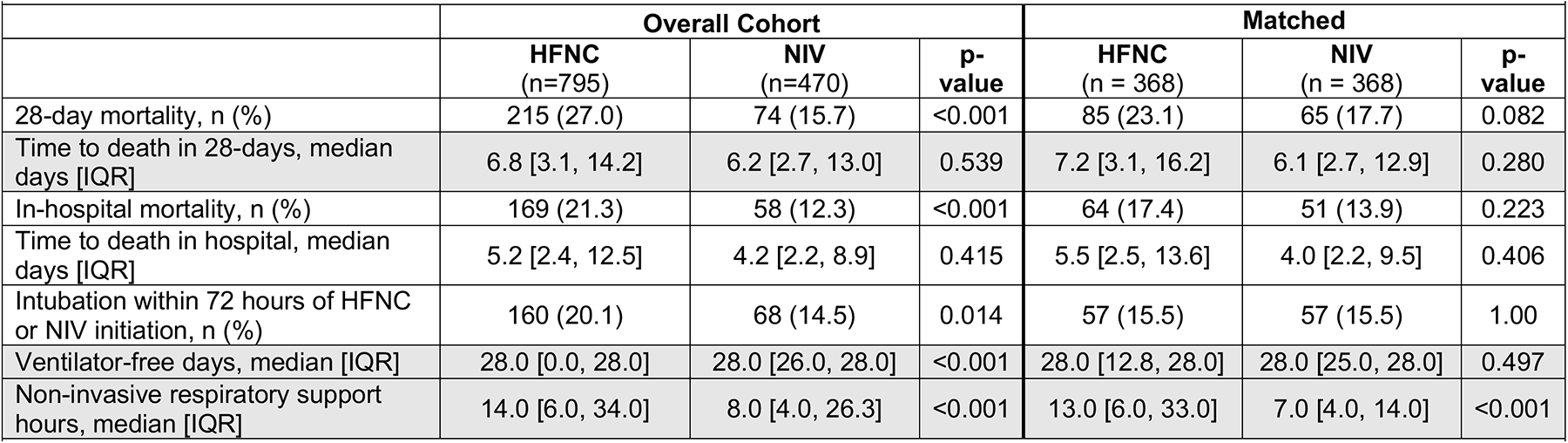
Individual patient outcomes before and after propensity score matching. Data are presented as n(%) and median [IQR]. p-values were calculated using Mann-Whitney U for continuous variables and Chi-squared test for categorical variables. Grey rows represent outcomes that contribute to the primary composite Major Adverse Pulmonary Events outcome. Ventilator-free days were calculated from admission through day 28. Non-invasive respiratory support hours were hours spent on HFNC or NIV calculated from initiation through hour 72. <u>Definitions</u>: HFNC= high flow nasal cannula, NIV= non-invasive positive pressure ventilation.

|  | Overall Cohort |  |  | Matched |  |  |
| --- | --- | --- | --- | --- | --- | --- |
|  | HFNC<br>(n=795) | NIV<br>(n=470) | p-value | HFNC<br>(n = 368) | NIV<br>(n = 368) | p-value |
| 28-day mortality, n (%) | 215 (27.0) | 74 (15.7) | <0.001 | 85 (23.1) | 65 (17.7) | 0.082 |
| Time to death in 28-days, median days [IQR] | 6.8 [3.1, 14.2] | 6.2 [2.7, 13.0] | 0.539 | 7.2 [3.1, 16.2] | 6.1 [2.7, 12.9] | 0.280 |
| In-hospital mortality, n (%) | 169 (21.3) | 58 (12.3) | <0.001 | 64 (17.4) | 51 (13.9) | 0.223 |
| Time to death in hospital, median days [IQR] | 5.2 [2.4, 12.5] | 4.2 [2.2, 8.9] | 0.415 | 5.5 [2.5, 13.6] | 4.0 [2.2, 9.5] | 0.406 |
| Intubation within 72 hours of HFNC or NIV initiation, n (%) | 160 (20.1) | 68 (14.5) | 0.014 | 57 (15.5) | 57 (15.5) | 1.00 |
| Ventilator-free days, median [IQR] | 28.0 [0.0, 28.0] | 28.0 [26.0, 28.0] | <0.001 | 28.0 [12.8, 28.0] | 28.0 [25.0, 28.0] | 0.497 |
| Non-invasive respiratory support hours, median [IQR] | 14.0 [6.0, 34.0] | 8.0 [4.0, 26.3] | <0.001 | 13.0 [6.0, 33.0] | 7.0 [4.0, 14.0] | <0.001 |

**Table 3.**
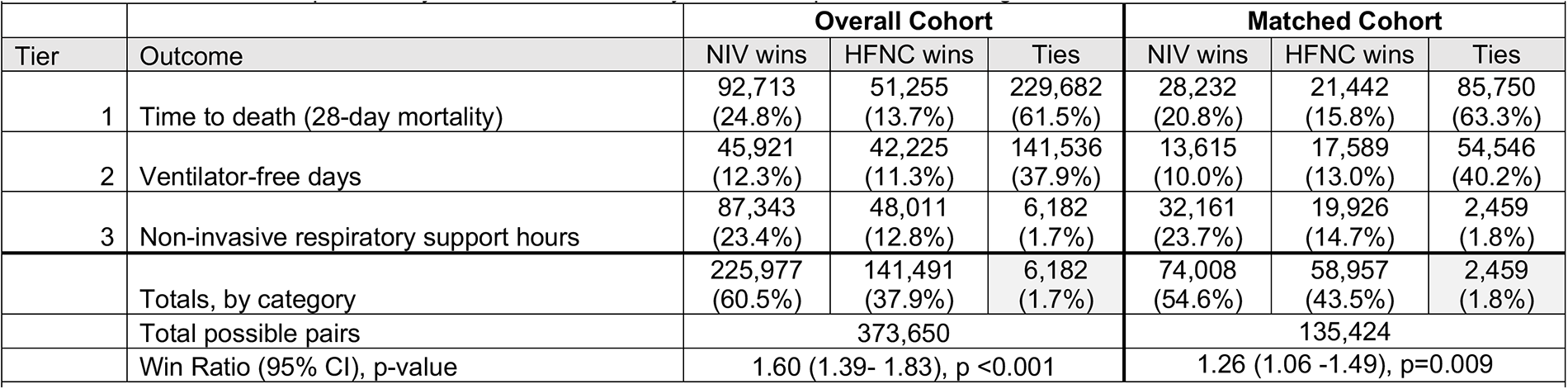
Details of the Win Ratio used to calculate the composite Major Adverse Pulmonary Events in the overall cohort (N=1,265) and in the matched study cohort (N=736). The Win Ratio in the matched cohort is the primary outcome, which is also displayed visually in Figure 2. The Win Ratio is the ratio of overall “wins for NIV” over “wins for HFNC.” A positive Win Ratio suggests NIV results in a better composite outcome compared to HFNC. Percentages represent the percent of pairs out of the total possible pairs. On tier 1 (time to death) and overall, the percentages sum to 100% because all patient pairs are compared at these levels. However, tiers 2 and 3 only compare patients who tied on the previous tier. For example, in tier 2 (ventilator-free days), the percentages add to the number of patients who tied on the previous tier. <u>Definitions:</u> HFNC= high flow nasal cannula, NIV= non-invasive positive pressure ventilation. Ventilator-free days were calculated from admission through day 28. Non-invasive respiratory support hours were hours spent on HFNC or NIV calculated from initiation through hour 72.

We then calculated the composite major adverse pulmonary events using the Win Ratio. There were 135,424 potential matched patient pairs (368x368). In total, NIV won in 74,008 (54.6%) pairs while HFNC won in 58,957 (43.5%), resulting in a Win Ratio for NIV of 1.26 (95% CI: 1.06-1.49, p=0.009). NIV won over HFNC for 28-day mortality (wins, as percent of all pairs: 20.8% vs 15.8%) and NIRS hours (wins: 23.7% vs 14.7%), but not ventilator-free days (wins: 10.1% vs 13.0%). Only 2,459 (1.8%) pairs tied on all tiers (**Table 2**, **Figure 2**) Results were robust to sensitivity analyses including patients with early respiratory mode cross-over and discontinuation (**eTable 3**) and alternative approaches to Win Ratio calculation (**eTable 4**).

**Figure 2.**
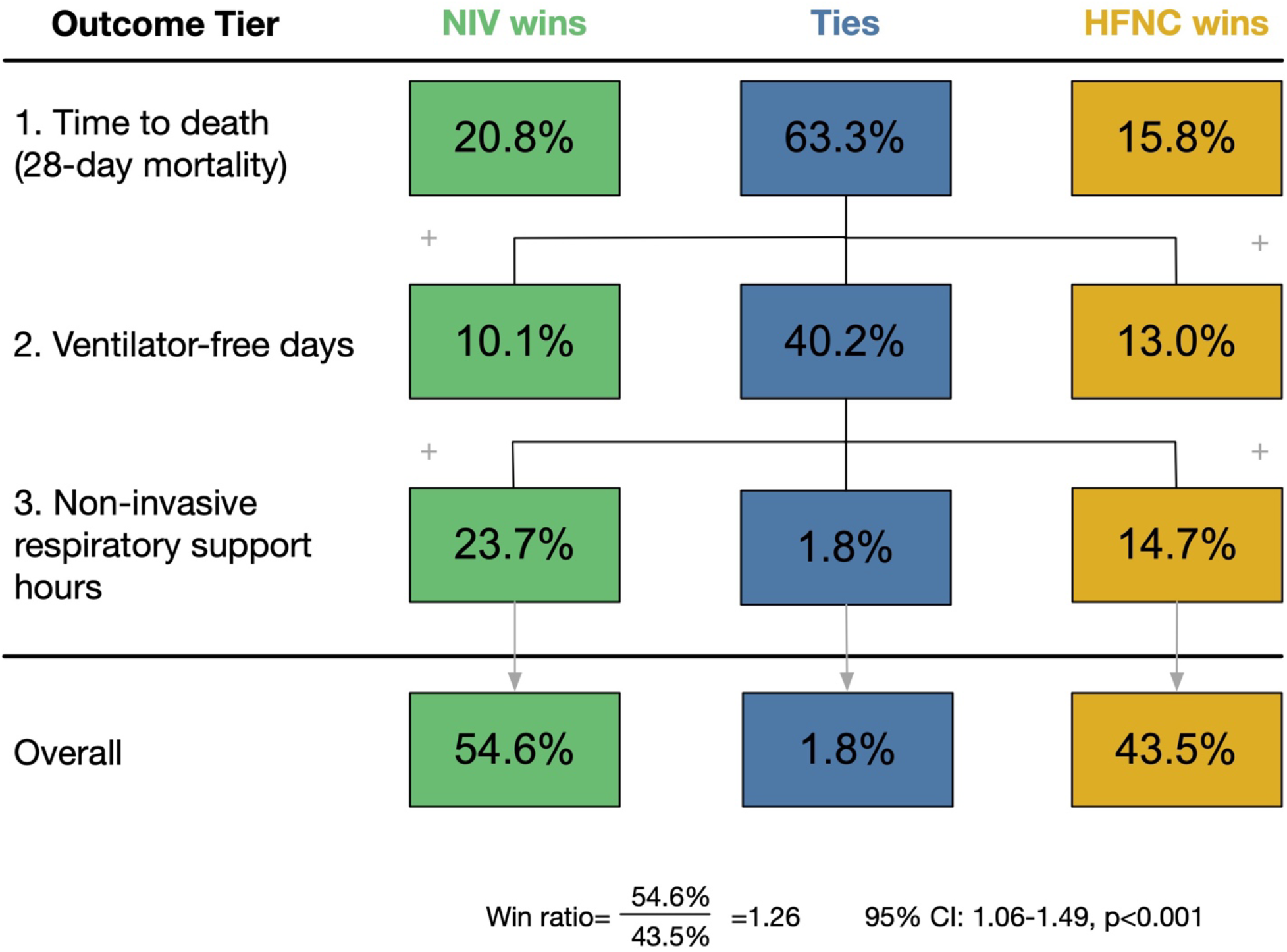
Comparison of Major Pulmonary Adverse Events between NIV and HFNC using the Win Ratio. The Win Ratio is the ratio of overall “wins for NIV” over “wins for HFNC.” A positive Win Ratio suggests NIV results in a better composite outcome compared to HFNC. Percentages represent the percent of pairs out of the total possible pairs. On tier 1 (time to death) and overall, the percentages sum to 100% because all patient pairs are compared at these levels. However, tiers 2 and 3 only compare patients who tied on the previous tier. For example, in tier 2 (ventilator-free days), the percentages add to the number of patients who tied on the previous tier (63.3%). <u>Definitions</u>: Non-invasive respiratory support hours = time spent on non-invasive respiratory support (NIV or HFNC) in hours, from initiation through hour 72. NIV= non-invasive ventilation. HFNC= high-flow nasal cannula.

## Discussion

In this propensity matched retrospective study of patients with acute hypoxemic respiratory failure, initial treatment with NIV decreased time spent on non-invasive respiratory support in the first 72 hours but did not have a significant impact on individual outcomes of mortality or ventilator-free days. However, treatment with NIV was associated with lower composite major adverse pulmonary events, calculated using a Win Ratio.

Our finding that initial treatment with NIV may decrease overall major adverse pulmonary events compared to HFNC contrasts with the findings of the prominent FLORALI trial. In that trial, HFNC improved mortality and ventilator-free days compared to NIV.^13^ Unfortunately, the results of the FLORALI trial have not been replicated consistently in the few other trials that have have compared HFNC and NIV in acute hypoxemic respiratory failure.^3,4,14,15,20^ The failure to replicate these results may be due to limitations of the FLORALI trial (small sample size with potentially high fragility index) and/or due to differences in patient populations enrolled across studies. For example, while our study included all patients receiving HFNC or NIV within 24 hours of ED arrival, the FLORALI trial enrolled a much narrower population of ICU patients with pure hypoxemic respiratory failure, excluding patients with hypercarbia, cardiogenic pulmonary edema, or COPD exacerbations.^13^ HFNC and NIV have not been directly compared in these other conditions (i.e.,heart failure and COPD exacerbations), where recommendations to use NIV are based on comparisons of NIV to low-flow oxygen not HFNC.^3,11^

There has only been one trial comparing HFNC vs NIV in a broad population of patients.^12^ Similar to our study, that trial by Doshi *et al* enrolled ED patients with acute hypoxemic respiratory failure and found HFNC was non-inferior to NIV based on intubation rates, though the trial was small and not powered to assess other outcomes. Therefore, the optimal non-invasive respiratory support mode for treating early, undifferentiated hypoxemic respiratory failure remains unclear. Our results add to clinical equipoise by suggesting that initial use of NIV may improve outcomes compared to HFNC in the broader population of patients with early, undifferentiated hypoxemic respiratory failure.

Understanding the optimal initial treatment for undifferentiated hypoxemic respiratory failure is critical, particularly in the ED, where the choice between HFNC and NIV often must be made before information about a patient’s diagnosis is fully available. Indeed, our findings suggest that in practice patients frequently have multiple risk factors for respiratory failure. For example, even among the patients who received HFNC, over half had a documented history of heart failure and/or COPD. While the frequency of these comorbidities may be high due to our institution’s role as a tertiary referral center, this finding suggests that it may be challenging to identify patients with specific etiologies of respiratory failure in real time. This is further reflected by the mix of encounter diagnosis codes in our cohort, which suggest many patients had mixed etiologies of respiratory failure.

Our results underscore the need for a randomized controlled trial to further understand the impact of NIV vs HFNC in patients with early, undifferentiated hypoxemic respiratory failure. In order to best inform practice decisions, such trials must employ novel design approaches, such as exemption from informed consent and pragmatic designs^21^, to capture patients early in their disease course, ideally in the ED at the time of NIV or HFNC initiaiton. Future clinical trials must also select appropriate outcomes to overcome the limitations of potentially subjective primary outcomes, such as intubation or ventilator-free days.

Our study suggests that use of a Win Ratio may be a feasible approach to using composite outcomes in future studies of respiratory support modes. In our study, calculating the composite outcome of major adverse pulmonary events using a Win Ratio allowed a more nuanced understanding of the impact of NIV and HFNC on patient outcomes than individual outcomes alone. At the individual outcome level, we found no association between respiratory support mode and mortality or ventilator-free days. In contrast, based on the composite outcome of major adverse pulmonary events calculated using a Win Ratio, NIV was superior to HFNC overall and at the mortality and respiratory support hour tiers. These findings highlight one challenge of studies of respiratory failure: the limitations of single outcomes such as mortality or intubation, which can obscure other clinically important outcomes particularly when only a subset of patients experience death or intubation. For example, in our study, a majority of patients survived and were never intubated, but patients treated initially with NIV spent less total time on non-invasive respiratory support. Given both HFNC and NIV require ICU level support at many institutions, spending less time on these devices is a potentially important outcome in patients who survive and are not intubated. Additionally, decisions about intubation and extubation are often subjective^16^, limiting the utility of intubation and ventilator-free days as primary outcomes. The Win Ratio offers a mechanism to compare multiple relevant patient outcomes, while maintaining a hierarchical approach that reflects the relative importance of outcomes to patients and their families: first comparing mortality, then ventilator-free days, then non-invasive respiratory support hours. Our results suggest that a similar Win Ratio could be used as a primary outcome for future clinical trials comparing respiratory support modalities.

### Limitations

This study has several limitations common to retrospective studies. First, there is a risk that our findings are the result of residual confounding. The use of propensity score matching limits that risk, but the presence of confounding on unmeasured variables remains possible and a prospective clinical trial is needed to verify these findings. Secondly, the assignment of the exposure group required that patients receive HFNC or NIV exclusively for the first two hours after initiation, which may limit generalizability to patients who switch support modalities or discontinue them within the first two hours. However, in sensitivity analyses where the exposure group was assigned by first modality, a similar result was observed. Finally, as a single-center study performed at a tertiary care hospital, generalizability may be limited as practice patterns regarding HFNC and NIV use may differ at other centers. While indications for HFNC and NIV selection may vary across institutions, propensity matching helps ensure that the patients included in our study were similar and might be candidates for either HFNC or NIV more broadly.

## Conclusion

In this propensity matched retrospective study of patients with early acute hypoxemic respiratory failure, initial treatment with NIV was superior to HFNC for major adverse pulmonary events, calculated using a Win Ratio. These results underscore the need for novel randomized controlled trials to definitively determine the merits of each non-invasive respiratory support strategy.

## Data Availability

All data produced in the present study are available upon reasonable request to the authors

## Acknowledgements

This material is the result of work supported with resources and the use of facilities at the Ann Arbor VA Medical Center.

## Authors’ contributions

ESM, IP, and CMF contributed to the conceptualization and design of the study, data analysis and interpretation, and drafting of this manuscript. MH, WJM, JMM, MAT, WL, HW contributed to the conceptualization of the study, data interpretation, and the writing of the manuscript, including substantive revisions. HCP contributed to data interpretation and the writing of the manuscript, including substantive revisions. All authors have read and approved the final manuscript.

## Impact

In this propensity score matched retrospective study of patients with early acute hypoxemic respiratory failure, initial treatment with non-invasive ventilation was superior to high-flow nasal cannula for major adverse pulmonary events, calculated using a Win Ratio. These results underscore the need for novel randomized controlled trials to definitively determine the merits of each non-invasive respiratory support strategy.

