## Supplemental Tables and Figures for "High-flow nasal cannula vs non-invasive ventilation in acute hypoxia: Propensity score matched study"

**Online Supplement: Table of Contents**

**eTables**
eTable 1: Variable definitions and sources

eTable 2: ICD-10 diagnosis codes by non-invasive respiratory support mode

eTable 3: Sensitivity analysis including patients with early cross-over between non-invasive respiratory support modalities and early discontinuation of HFNC or NIV

eTable 4: Sensitivity analysis: Alternative approaches to Win Ratio calculations

**eFigures**
eFigure 1. Logistic regression model used to calculate the propensity score for odds of treatment with non-invasive ventilation

eFigure 2. Covariate balance after propensity score matching

eFigure 3. Hourly respiratory support from non-invasive respiratory support initiation through hour 72, in the overall and matched cohorts

eFigure 4. Daily status plot from emergency department arrival through day 28, in the overall and matched cohorts

eFigure 5. Kaplan-Meier survival curves for 28-day survival and time to intubation, in the overall and matched cohorts

| **eTable 1.** Variable definitions and sources | | |
|---|---|---|
| **Variable** | **Definition** | **Source** |
| Adult ED encounter | Any hospital encounter beginning in the adult emergency department | Clarity - ED Datamart |
| 6 L/min or more oxygen flow rate in ED | Any flowsheet recording of 6L/min of oxygen flow or higher via any respiratory support device prior to leaving ED | Caboodle - Flowsheets |
| Mechanical vent w/o NIRS | Receipt of invasive mechanical ventilation without first having received NIRS | Caboodle - Flowsheets |
| NIRS in first 24 hours | Any heated high-flow nasal cannula or non-invasive ventilation support in first 24 hours from ED arrival | Caboodle - Flowsheets |
| Chronic assisted ventilation | Any encounter with respiratory therapy documentation for chronic assisted ventilation or ventilator mode used for chronic assisted ventilation at our institution (AVAPS). | Clarity - ED Datamart |
| Tracheostomy patient | Any encounter with flowsheet (lines/drains/airways or tracheostmy care) documentation during first hospital day | Caboodle - Flowsheets |
| NIRS post-extubation only | Any occurrence of NIRS documentation that occurred only after invasive mechanical ventilation | Caboodle - Flowsheets |
| Covid-19 positive | Covid-19 positive by institutional guidelines (positive test or physician's documentation of an outside positive test within 21 days prior to start of encounter) | Clarity - ED Datamart |
| Age | Age in years at the start of the encounter | Clarity - ED Datamart |
| Sex at birth | Sex at birth | Clarity - ED Datamart |
| Body mass index | Weight in kg divided by cm2 using the median weight of the encounter | Clarity - ED Datamart |
| Charlson comorbidity index | Standard Charlson Comorbidity Index using the technique described in Quan et al. (Am J Epi 2011) | Clarity - ED Datamart |
| Congestive heart failure | ICD-10 codes: I42.x, I43.x, I50.x | Clarity - ED Datamart |
| Chronic obstructive pulmonary disease | ICD-10 codes: J40.x-J47.x, J60.x-J67.x, J68.4, J70.1, J70.3 | Clarity - ED Datamart |
| Obstructive sleep apnea | ICD-10 codes: G47.3 | Clarity - ED Datamart |
| Expected $pCO_2$ | Calculated using the first available bicarbonate from basic metabolic panel and pCO2 from arterial or venous blood gas, comparing actual $pCO_2$ to $pCO_2$ that would be expected based on Winter's formula | Clarity - ED Datamart |
| First hour of NIRS | First hour of any NIRS, either HFNC or NIV | Caboodle - Flowsheets |
| Systolic shock index | Heart rate (beats per minute) divided by systolic blood pressure (mmHg) | Clarity - ED Datamart |
| Lactate | Serum lactate measured by any means (blood gas, separate lab assay, or point-of-care) | Clarity - ED Datamart |
| Glasgow coma scale | Standard total GCS obtained by nursing | Caboodle - Flowsheets |
| Highest day 1 SOFA | Highest total sequential organ failure assessment score measured in the first 24 hours from ED arrival | Clarity - ED Datamart |
| 28-day mortality | Deceased at the end of the 28th day from ED arrival. Includes out of hospital deaths after discharge. | Clarity - ED Datamart |

| | | |
|---|---|---|
| Time to death (28-days) | Time in days from ED arrival to death censored at 28 days | Clarity - ED Datamart |
| In-hospital mortality | Deceased at discharge from hospital | Clarity - ED Datamart |
| Time to death (in-hospital) | Time in days from ED arrival to death censored at hospital discharge | Clarity - ED Datamart |
| Ventilator-free days | Number of days free of invasive mechanical ventilation in the first 28 days from ED arrival. If deceased within 28 days then ventilator-free days = 0. | Clarity - ED Datamart |
| Non-invasive respiratory support hours | Total hours of either NIV or HFNC from initiation through hour 72. | Caboodle - Flowsheets |
| Sepsis | ICD-10 codes: A02.1, A20.7, A22.7, A26.7, A32.7, A39.2, A39.3, A39.4, A40.0, A40.1, A40.3, A40.8, A40.9, A41.01, A41.02, A41.1, A41.2, A41.3, A41.4, A41.50, A41.51, A41.52, A41.53, A41.59, A41.81, A41.89, A41.9, A42.7, A54.86, B00.7, B37.7, I76., O03.37, O03.87, O04.87, O07.37, O08.82, O85., O86.04, P36.0, P36.10, P36.19, P36.2, P36.30, P36.39, P36.4, P36.5, P36.8, P36.9, R65.20, R65.21, T81.12XA, T81.44XA | Clarity - ED Datamart |
| Pneumonia | ICD-10 codes: A01.03, A02.22, A20.2, A21.2, A22.1, A31.0, A37.01, A37.11, A37.81, A37.91, A43.0, A48.1, A50.04, A52.72, A54.84, B01.2, B05.2, B06.81, B25.0, B37.1, B38.0, B38.1, B38.2, B39.0, B39.1, B39.2, B58.3, B59., B77.81, J09.X1, J10.00, J10.01, J10.08, J11.00, J11.08, J12.0, J12.1, J12.2, J12.3, J12.81, J12.82, J12.89, J12.9, J13., J14., J15.0, J15.1, J15.20, J15.211, J15.212, J15.29, J15.3, J15.4, J15.5, J15.6, J15.7, J15.8, J15.9, J16.0, J16.8, J17., J18.0, J18.1, J18.8, J18.9, J85.1, J95.851, J09.X1, J09.X2, J09.X3, J09.X9, J10.00, J10.01, J10.08, J10.1, J10.2, J10.81, J10.82, J10.83, J10.89, J11.00, J11.08, J11.1, J11.2, J11.81, J11.82, J11.83, J11.89, J45.20, J45.21, J45.22, J45.30, J45.31, J45.32, J45.40, J45.41, J45.42, J45.50, J45.51, J45.52, J45.901, J45.902, J45.909, J45.990, J45.991, J45.998 | Clarity - ED Datamart |
| Aspiration | ICD-10 codes: J69.0, J69.1, J69.8, J95.4, O29.011, O29.012, O29.013, O29.019, O74.0, O89.01 | Clarity - ED Datamart |
| ARDS | ICD-10 codes: J80 | Clarity - ED Datamart |
| Hypercarbic respiratory failure | ICD-10 codes: J96.02, J96.22, J96.92, J96.12 | Clarity - ED Datamart |
| Respiratory failure, not otherwise specified | ICD-10 codes: J95.821, J95.822, J96.00, J96.01, J96.20, J96.21, J96.90, J96.91, R0.92, J96.10, J96.11 | Clarity - ED Datamart |
| Chronic obstructive pulmonary disease or asthma | ICD-10 codes: J41.0, J41.1, J41.8, J42x, J43.0, J43.1, J43.2, J43.8, J43.9, J44.0, J44.1, J44.9, J45.20, J45.21, J45.22, J45.30,J45.31, J45.32, J45.40, J45.41, J45.42, J45.50, J45.51, J45.52, J459.01, J45.902, J45.909, J45.990, J45.991, J45.998 | Clarity - ED Datamart |
| Volume overload | ICD-10 codes: J81.0, I12.0, I12.9, I13.0, I13.10, I13.11, I13.2, E87.70, E87.71, E87.79, I09.81, I11.0, I13.0, I13.2, I50.1, I50.20, I50.21, I50.22, I50.23, I50.30, I50.31, I50.32, I50.33, I50.40, I50.41, I50.42, I50.43, I50.810, I50.811, I50.812, I50.813, I50.814, I50.82, I50.83, I50.84, I50.89, I50.9, I97.130, I97.131, O29.121, O29.122, O29.123, O29.129, I11.0, I11.9, I12.0, I12.9, I13.0, I13.10, I13.11, I13.2, I15.0, I15.1, I15.2, I15.8, I15.9, I16.0, I16.1, I16.9 | Clarity - ED Datamart |

**eTable 2.** ICD-10 diagnosis codes, by non-invasive respiratory support mode

| | **Overall Cohort** | | **Matched Cohort** | |
|---|---|---|---|---|
| | **HFNC** (n=795) | **NIV** (n=470) | **HFNC** (n = 368) | **NIV** (n = 368) |
| Sepsis | 262 (32.9%) | 86 (18.3%) | 101 (27.4%) | 76 (20.7%) |
| Pneumonia | 389 (48.9%) | 183 (38.9%) | 171 (46.5%) | 151 (41.0%) |
| Aspiration | 129 (16.2%) | 28 (6.0%) | 45 (12.2%) | 23 (6.3%) |
| Acute respiratory distress syndrome | 28 (3.5%) | 8 (1.7%) | 10 (2.7%) | 7 (1.9%) |
| Hypercarbic respiratory failure | 111 (14.0%) | 226 (48.1%) | 75 (20.4%) | 159 (43.2%) |
| Respiratory failure, not otherwise specified | 713 (89.7%) | 402 (85.5%) | 333 (90.5%) | 313 (85.1%) |
| Chronic obstructive pulmonary disease or asthma | 276 (34.7%) | 246 (52.3%) | 150 (40.8%) | 184 (50.0%) |
| Volume overload | 430 (54.1%) | 367 (78.1%) | 239 (64.9%) | 276 (75.0%) |

**eTable 2 Legend.** ICD-10 admission codes recorded for the encounter. All respiratory-related ICD-10 codes for each encounter were included, so one encounter could be associated with multiple diagnosis codes. Diagnosis codes were grouped using the Clinical Classifications Software Refined (CCSR), which is made available through the Agency for Healthcare Research and Quality. Accessed 8/17/2023 via https://hcup-us.ahrq.gov/toolssoftware/ccsr/ccs_refined.jsp. Volume overload included diagnoses of volume overload in setting of heart failure, kidney failure, and / or hypertensive emergency.

Definitions: HFNC= high flow nasal cannula, NIV= non-invasive positive pressure ventilation

**eTable 3:** Sensitivity analysis: Win Ratio based on first non-invasive respiratory support mode

| | | Overall expanded cohort | | | Matched expanded cohort | | |
|---|---|---|---|---|---|---|---|
| **Tiers** | **Outcome** | **NIV wins** | **HFNC wins** | **Ties** | **NIV wins** | **HFNC wins** | **Ties** |
| 1 | Time to death (28-day mortality) | 143,064 (23.4%) | 89,040 (14.6%) | 378,230 (62.0%) | 52,535 (21.1%) | 37,903 (15.2%) | 158,563 (63.7%) |
| 2 | Ventilator-free days | 76,426 (12.5%) | 69,931 (11.5%) | 231,873 (40.0%) | 28,010 (11.2%) | 30,360 (12.2%) | 100,193 (40.2%) |
| 3 | Non-invasive respiratory support hours | 128,786 (21.1%) | 92,457 (15.1%) | 10,630 (1.7%) | 53,697 (21.6%) | 41,745 (16.8%) | 4,751 (1.9%) |
| | Totals, by category | 348,276 (57.1%) | 251,428 (41.2%) | 10,630 (1.7%) | 134,242 (53.9%) | 110,008 (44.2%) | 4,751 (1.9%) |
| | Total possible pairs | 610,334 | | | 249,001 | | |
| | Win Ratio (95% CI), p-value | 1.39 (1.23 – 1.56), p-value <0.001 | | | 1.22 (1.05 – 1.41), p-value= 0.0079 | | |

**eTable 3 Legend.**
We performed a sensitivity analysis classifying patients as receiving NIV vs HFNC based on first respiratory support mode, to mimic an intention-to-treat approach. In this analysis, we included patients with early non-invasive respiratory support discontinuation and cross-over (See figure 1). In the dataset, the hourly non-invasive respiratory support mode is the dominant mode (mode used most frequently during the hour). Of 1,615 patients with qualifying non-invasive respiratory support mode for inclusion in this sensitivity analysis, 10 patients received NIV and HFNC equally in hour 1 and a dominant mode could not be determined. These patients were therefore excluded, leaving N=1,595. Using a propensity-score for odds of treatment with NIV, we were able to match 998/1,595 encounters (62.6%: HFNC N=499, NIV N=499). We calculated the Win Ratio for composite Major Adverse Pulmonary Events in both the overall expanded cohort and the expanded cohort after propensity score matching. The Win Ratio is the ratio of overall "wins for NIV" over "wins for HFNC." A positive Win Ratio suggests NIV results in a better composite outcome than HFNC. See methods, Table 3 and Figure 2 for full descriptions of Win Ratio calculation.

Definitions: HFNC= high flow nasal cannula, NIV= non-invasive positive pressure ventilation, NIRS= non-invasive respiratory support (either HFNC or NIV). Non-invasive respiratory support hours were hours spent on HFNC or NIV calculated from initiation through hour 72.

**eTable 4.** Sensitivity analysis: Alternative approaches to Win Ratio calculations

| | | Overall Cohort | | | Matched Cohort | | |
|---|---|---|---|---|---|---|---|
| | | 373,650 total pairs | | | 135,424 total pairs | | |
| **A. 28-day mortality, as a survival event (Primary analysis, corresponds to Figure 2)** | | | | | | | |
| Tier | Outcome | NIV wins | HFNC wins | Ties | NIV wins | HFNC wins | Ties |
| 1 | Time to death (28 day mortality) | 92,713 (24.8%) | 51,255 (13.7%) | 229,682 (61.5%) | 28,232 (20.8%) | 21,442 (15.8%) | 85,750 (63.3%) |
| 2 | Ventilator-free days | 45,921 (12.3%) | 42,225 (11.3%) | 141,536 (37.9%) | 13,615 (10.0%) | 17,589 (13.0%) | 54,546 (40.2%) |
| 3 | Non-invasive respiratory support hours | 87,343 (23.4%) | 48,011 (12.8%) | 6,182 (1.7%) | 32,161 (23.7%) | 19,926 (14.7%) | 2,459 (1.8%) |
| | Totals, by category | 225,977 (60.5%) | 141,491 (37.9%) | 6,182 (1.7%) | 74,008 (54.6%) | 58,957 (43.5%) | 2,459 (1.8%) |
| | Win Ratio (95% CI), p-value | 1.60 (1.39- 1.83), p <0.0001 | | | 1.26 (1.06 -1.49), p=0.0092 | | |
| **B. 28-day mortality, as a binary event** | | | | | | | |
| Tier | Outcome | NIV wins | HFNC wins | Ties | NIV wins | HFNC wins | Ties |
| 1 | 28-day mortality | 85,140 (22.8%) | 42,920 (11.5%) | 245,590 (65.7%) | 25,755 (19.0%) | 18,395 (13.6%) | 91,274 (67.4%) |
| 2 | Ventilator-free days | 45,921 (12.3%) | 42,225 (11.3%) | 157,444 (42.1%) | 13,615 (10.1%) | 17,589 (13.0%) | 60,070 (44.4%) |
| 3 | Non-invasive respiratory support hours | 87,343 (23.4%) | 48,011 (12.8%) | 22,090 (5.9%) | 32,161 (23.7%) | 19,926 (14.7%) | 7,983 (5.9%) |
| | Totals, by category | 218,404 (58.5%) | 133,156 (35.6%) | 22,090 (5.9%) | 71,531 (52.8%) | 55,910 (41.3%) | 7,983 (5.9%) |
| | Win Ratio (95% CI), p-value | 1.64 (1.42-1.89), p <0.0001 | | | 1.28 (1.07-1.53), p=0.0066 | | |
| **C. In-hospital mortality, as a survival event** | | | | | | | |
| Tier | Outcome | NIV wins | HFNC wins | Ties | NIV wins | HFNC wins | Ties |
| 1 | Time to death (In-hospital mortality) | 44,200 (11.8%) | 29,335 (7.9%) | 300,115 (80.3%) | 12,835 (9.5%) | 11,927 (8.8%) | 110,662 (81.7%) |
| 2 | Ventilator-free days | 93,374 (25.0%) | 62,052 (16.6%) | 144,689 (38.7%) | 28,896 (21.3%) | 26,182 (19.3%) | 55,584 (41.0%) |

| | | | | | | | |
|---|---|---|---|---|---|---|---|
| 3 | Non-invasive respiratory support hours | 88,774 (23.8%) | 49,636 (13.3%) | 6,279 (1.7%) | 32,590 (24.1%) | 20,506 (15.1%) | 2,488 (1.8%) |
| | Totals, by category | 226,348 (60.6%) | 141,023 (37.7%) | 6,279 (1.7%) | 74,321 (54.9%) | 58,615 (43.3%) | 2,488 (1.8%) |
| | Win Ratio (95% CI), p-value | 1.61 (1.40- 1.84), p <0.0001 | | | 1.27 (1.07 -1.50), p=0.0066 | | |
| **D. In-hospital mortality as a binary event** | | | | | | | |
| Tier | Outcome | NIV wins | HFNC wins | Ties | NIV wins | HFNC wins | Ties |
| 1 | In-hospital mortality | 69,628 (18.6%) | 36,308 (9.7%) | 267,714 (71.6%) | 20,288 (15.0%) | 15,504 (11.4%) | 99,632 (73.6%) |
| 2 | Ventilator-free days | 64,793 (17.3%) | 51,202 (13.7%) | 151,719 (40.6%) | 20,467 (15.1%) | 21,360 (15.8%) | 57,805 (42.7%) |
| 3 | Non-invasive respiratory support hours | 87,462 (23.4%) | 48,261 (36.3%) | 15,996 (4.3%) | 32,115 (23.7%) | 19,972 (14.7%) | 5,718 (4.2%) |
| | Totals, by category | 221,883 (59.4%) | 135,771 (36.3%) | 15,996 (4.3%) | 72,870 (53.8%) | 56,836 (42.0%) | 5718 (4.2%) |
| | Win Ratio (95% CI), p-value | 1.63 (1.42- 1.88), p <0.0001 | | | 1.28 (1.08 -1.53), p=0.0054 | | |

**eTable 4 Legend.** Details of alternative approaches used to calculate the Win Ratio for composite Major Adverse Pulmonary Events.

In the primary analysis, 28-day mortality was used as a survival event (Panel A above, Table 3, Figure 2). Mortality as a survival event was chosen given this has been the convention in existing literature on the use of Win Ratio.[17] However, given the clinical significance of longer survival within 28-days is less clear in critical care, in sensitivity analysis, we also calculated the Win Ratio for 28-day mortality as a binary event (Panel B). In this scenario, patients win on mortality only if they lived but their pair died; time to death is not compared. Patients who both died tied on all levels and did not contribute to the final Win Ratio outcome. This approach results in differences on Tier 1 as well as an increased number of total ties. Sensitivity analyses were also performed using in-hospital mortality as both a C) survival and D) binary event.

Given the novelty of the Win Ratio as an outcome, as an exploratory analysis, we calculated the Win Ratio for the overall cohort (N=1,265) and the matched cohort (N=736). Interpretation of the results for the overall cohort is limited given the differences between baseline groups.

Definitions: HFNC= high flow nasal cannula, NIV= non-invasive positive pressure ventilation, NIRS= non-invasive respiratory support (either HFNC or NIV). Non-invasive respiratory support hours were hours spent on HFNC or NIV calculated from initiation through hour 72.

**eFigure 1**: Logistic regression model used to calculate the propensity score for odds of treatment with non-invasive ventilation

| Variable | | N | Odds ratio | | p |
|---|---|---|---|---|---|
| **Age, years** | | 1265 | | 1.01 (1.00, 1.02) | 0.080 |
| **Gender** | F | 554 | | Reference | |
| | M | 711 | | 0.91 (0.70, 1.19) | 0.500 |
| **BMI** | | 1265 | | 1.03 (1.02, 1.05) | <0.001 |
| **Charlson Index, Total** | | 1265 | | 1.02 (0.99, 1.04) | 0.143 |
| **CHF** | No | 464 | | Reference | |
| | Yes | 801 | | 2.29 (1.70, 3.11) | <0.001 |
| **COPD** | No | 457 | | Reference | |
| | Yes | 808 | | 0.95 (0.71, 1.27) | 0.722 |
| **OSA** | No | 973 | | Reference | |
| | Yes | 292 | | 1.02 (0.74, 1.40) | 0.900 |
| **Winter's** | Expected PCO2 | 254 | | Reference | |
| | PCO2 above expected | 586 | | 3.49 (2.48, 4.96) | <0.001 |
| | PCO2 below expected | 425 | | 0.74 (0.50, 1.09) | 0.128 |
| **Time to first NIRS, hours** | | 1265 | | 0.96 (0.94, 0.99) | 0.012 |
| **Systolic Shock Index** | | 1265 | | 0.46 (0.26, 0.82) | 0.009 |
| **Lactate** | Less than 2 mmol/L | 708 | | Reference | |
| | Between 2 and 4 | 392 | | 0.82 (0.61, 1.11) | 0.200 |
| | Greater than 4 | 165 | | 0.61 (0.39, 0.94) | 0.027 |
| **GCS** | | 1265 | | 1.08 (0.99, 1.19) | 0.098 |
| **Highest SOFA on day 1** | | 1265 | | 0.91 (0.86, 0.96) | <0.001 |

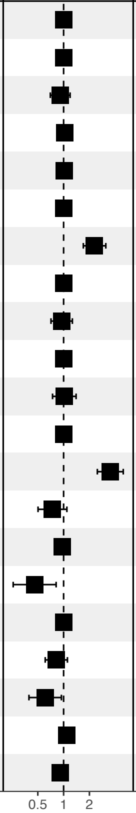


**eFigure 1 Legend**: Logistic regression model used to determine the propensity score for odds of receiving non-invasive ventilation (NIV). N=1,265 patients who met study inclusion criteria and were included in the pool for matching. Odds ratio represents odds of receiving NIV.

Definitions: BMI= body mass index; F= female; M= male; CHF= history of congestive heart failure; COPD= history of chronic obstructive pulmonary disease; OSA= history of obstructed sleep apnea; Expected $pCO_2$ at, above, or below expected = a comparison of actual $pCO_2$ on blood gas (venous or arterial) with the expected value as calculated using Winter's Formula and first bicarbonate value off of chemistry panel; Time to first NIRS, hours= hours from hospital arrival to initiation of non-invasive respiratory support (either high-flow nasal canula or non-invasive ventilation); Systolic shock index= ratio of heart rate / systolic blood pressure; GCS= Glasgow Coma Score; SOFA= Sequential Organ Failure Assessment.

**eFigure 2.** Covariate balance after propensity-score matching

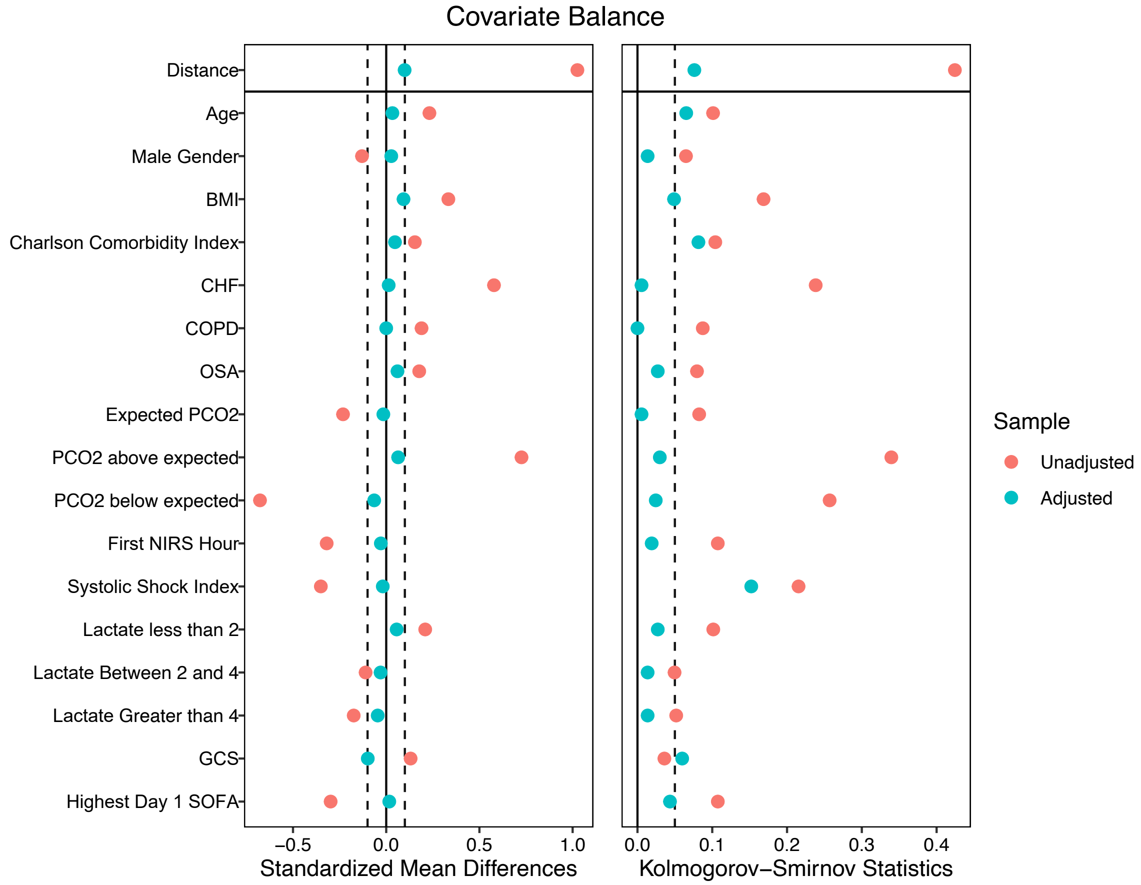


**eFigure 2 Legend**: Covariate balance after propensity score matching, displayed as both standardized mean differences and using Kolmogorov-Smirnov statistics. N=736 patients included in the matched cohort (368 in each group)

Definitions: BMI= body mass index; CHF= history of congestive heart failure; COPD= history of chronic obstructive pulmonary disease; OSA= history of obstructed sleep apnea; Expected $pCO_2$ at, above, or below expected = a comparison of actual $pCO_2$ on blood gas (venous or arterial) with the expected value as calculated using Winter's Formula and first bicarbonate value off of chemistry panel; First NIRS hour= time from hospital arrival to initiation of non-invasive respiratory support (either high flow nasal canula or non-invasive ventilation); Systolic shock index= ratio of heart rate / systolic blood pressure; GCS= Glasgow Coma Score; SOFA= Sequential Organ Failure Assessment.

**eFigure 3**. Hourly respiratory support from NIRS start through hour 72, in the overall and matched cohorts

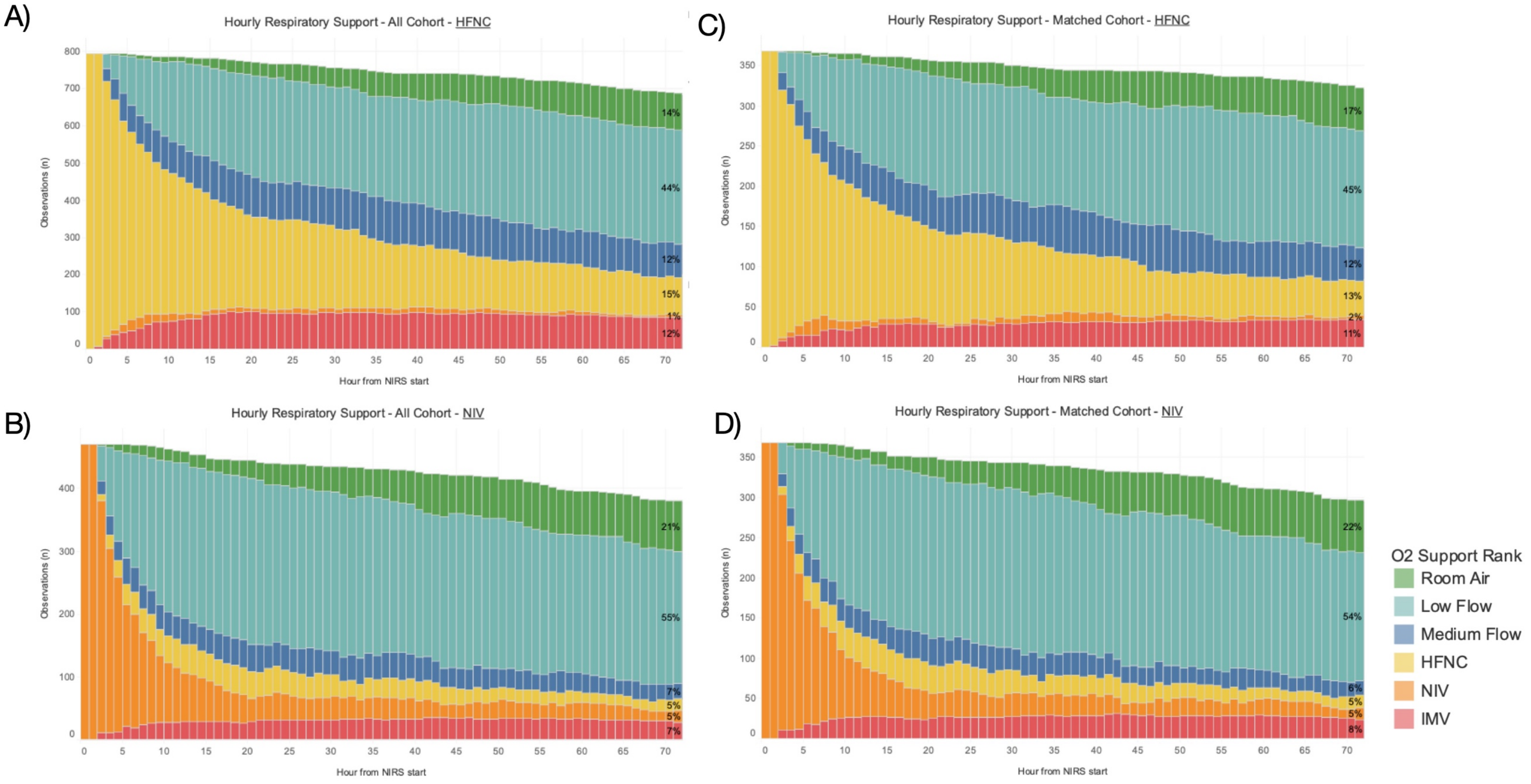


**eFigure 3 Legend**:
Respiratory support mode by hour, from initiation of non-invasive respiratory support (NIRS) through hour 72. Panels A and B display the hourly respiratory support modes for the overall cohort receiving A) HFNC (N=795) and B) NIV (N=407). Panels C and D represent only the matched cohort receiving C) HFNC (N=368) and D) NIV (N=368). Respiratory support modes are represented by colors, as detailed in the legend. The blank white space at the top of each graph represents patients who were discharged or died.

Definitions: Low flow oxygen= 1 to 6 Liters nasal canula; Medium flow oxygen= 6 to 15 liters nasal canula or oxygen mask; HFNC= high-flow nasal canula; NIV= non-invasive ventilation, which includes continuous positive airway pressure and bilevel positive airway pressure; IMV= invasive mechanical ventilation

**eFigure 4.** Daily status plot from ED arrival through day 28 in the overall and matched cohorts

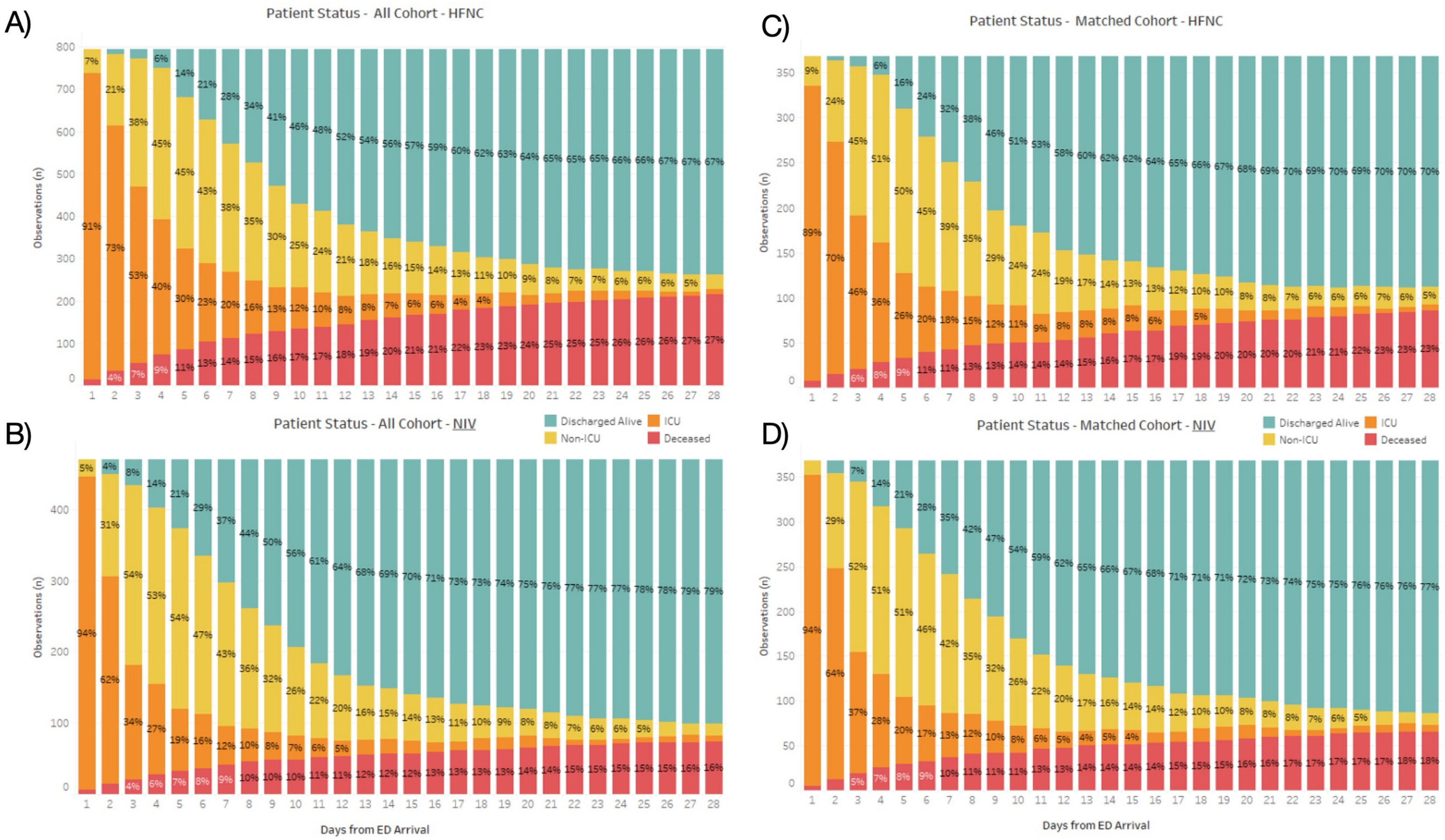


**eFigure 4 Legend**: Daily status plot from emergency department (ED) arrival through day 28. Panels A and B represent overall cohort receiving A) HFNC (N=795) and B) NIV (N=407). Panels C and D represent only the matched cohort receiving C) HFNC (N=368) and D) NIV (N=368).

Definitions: HFNC= high-flow nasal canula; NIV= non-invasive ventilation, which includes continuous positive airway pressure and bilevel positive airway pressure.

**eFigure 5.** Kaplan Meier survival curves for 28-day survival and time to intubation in the overall and matched cohorts.

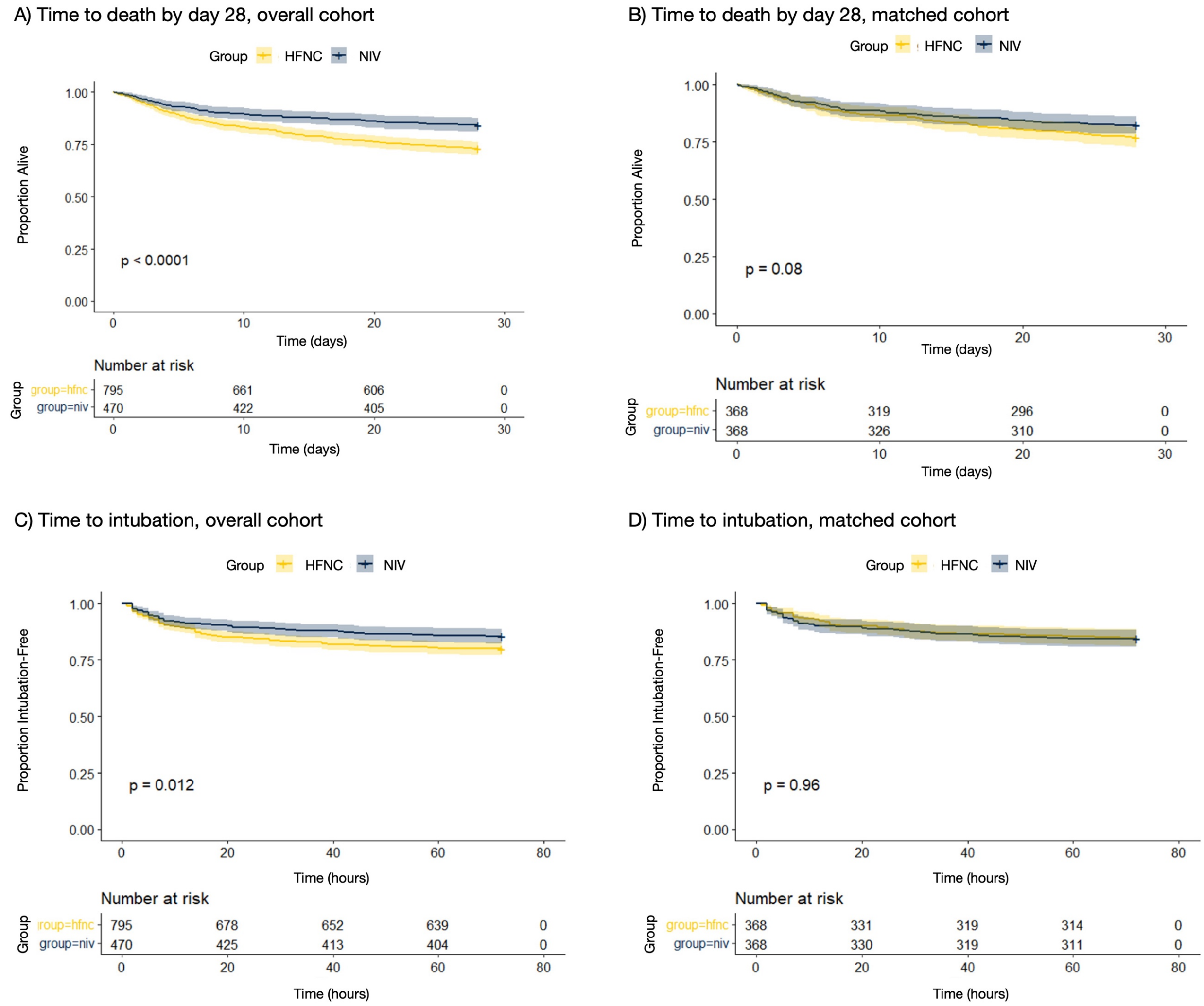


### eFigure 5 Legend

Top row: Kaplan-Meier 28-day survival curves for A) overall cohort (N=1,265) and B) matched cohort (N=736).

Bottom row: Kaplan-Meier survival curves for time to intubation, censored at 72 hours from the initiation of non-invasive respiratory support in C) overall cohort (N=1,265) and D) matched cohort (N=736).

Definitions: HFNC= high-flow nasal canula (represented in yellow); NIV= non-invasive ventilation, which includes continuous positive airway pressure and bilevel positive airway pressure (represented in blue).
